# Re-estimating prevalence of hepatitis B virus immunity among adults in the United States: self-reported vaccination, immunologic markers, and bias correction

**DOI:** 10.1101/2021.07.20.21260652

**Authors:** Daniel T. Vader, Chari Cohen, Neal D. Goldstein, Brian K. Lee, Harrison Quick, Alison A. Evans

## Abstract

**Background:** Two instruments used to measure adult hepatitis B vaccination coverage in the United States are self-report and antibody to hepatitis B surface antigen (anti-HBs). Estimates based on either of these measures are subject to misclassification when used to determine immunity to hepatitis B. This study presents misclassification-corrected estimates of hepatitis B immune prevalence in the US and compares them to self-report- and antibody-based estimates.

**Methods:** We used cross-sectional data from the 2015-2016 NHANES cycle on 5,151 adults in the US age 18 and older. Existing literature on long-term immunity after vaccination informed anti-HBs sensitivity as a measure of immunity. Our model incorporated literature-based distributions for sensitivity and specificity using a Bayesian approach to correct for misclassification of true immune status by anti-HBs.

**Results:** After correcting for misclassification, overall adult immune prevalence was estimated at 31.0% (95% credible interval, 27.9% to 34.1%). Anti-HBs prevalence was 6.4 (3.9 to 8.8) and self-report prevalence 2.6 (−0.6 to 5.8) percentage points lower than overall immune prevalence. Among Asian Americans, anti-HBs and self-report underestimated immune prevalence by 15.8% (11.5% to 20.9%) and 25.1% (17.3% to 33.2%), respectively. Among 19 to 25-year-olds, anti-HBs and self-report underestimated immune prevalence by 26.5% (20.7% to 32.5%) and 21.0% (12.6% to 28.9%).

**Conclusions:** Both self-reported vaccination and antibody-based measures underestimate hepatitis B immunity among adults. This underestimation was especially large among younger adults and Asian Americans. The consequences of treating these surrogates as unbiased measures of vaccination or immunity may only increase as more vaccinated children age into adulthood.

## Introduction

The success of hepatitis B prevention efforts in the United States over the past 30 years is – in large part – defined by development of an effective hepatitis B virus (HBV) vaccine and successful deployment of a national childhood vaccination strategy. The 3-dose hepatitis B vaccine series has an estimated 90% efficacy when administered to healthy adults and >95% efficacy when administered to individuals 18 years of age and younger.^1^ By 2004, following the introduction of a universal childhood vaccination strategy against hepatitis B in 1991, incidence of infection in the United States dropped by 75%.^2, 3^

But while hepatitis B vaccination coverage among children has been high since the 1990s, coverage among adults has remained low (likely due to child-focused vaccination strategy^4^). In 2016, an estimated 90.5% (95% CI 89.3%, 91.5%) of children aged 19 to 35 months were up-to-date on the hepatitis B vaccine series while only an estimated 24.8% (24.0%, 25.7%) of adults were up-to-date that same year.^5, 6^

However, there is more uncertainty surrounding the estimates of adult vaccination coverage compared to estimates of childhood coverage. While national estimates of HBV vaccination coverage among individuals age 17 and younger are based on provider records,^5, 7^ estimates of national adult vaccination coverage are reliant on the ability and willingness of individuals to correctly report their own vaccination status. The statistic for adult vaccination coverage cited above is based on data from the National Health Interview Survey, which relies exclusively on self-report to determine vaccination status.^6^ Rather than say that 24.8% of adults are fully vaccinated against HBV, it would be more accurate to say that, if asked, an estimated 24.8% of adults would report having received at least 3 doses of hepatitis B vaccine. In a separate section of the report that lists this statistic, the National Center for Immunization and Respiratory Diseases (NCIRD) admits that their official estimates may underestimate hepatitis B vaccination among young adults. As an example, they contend that self-report based statistics put vaccination coverage among 27-year-olds at 38.6% when coverage should be closer to 77% given historic data on the cohort.^6^ Based on these statistics, we suspect the sensitivity of self-report to be low (many people fail to self-report vaccination if they are truly vaccinated).

One alternative to self-report as a measure of vaccination is serological testing for markers of HBV immunity. Though testing requires the collection and examination of serum samples – which is more expensive and invasive than a survey – it is appealing because it does not rely on individual recall. However, serological tests are not perfect measures of vaccination status or even immunity to HBV.^8–10^ A key marker of HBV immunity that is used in serological testing is antibody to hepatitis B surface antigen (anti-HBs). In the years following vaccination, many individuals drop to low or undetectable levels of anti-HBs despite maintaining immune memory.^10,11,20–25,12–19^ If a previously vaccinated person with low or undetectable levels of anti-HBs is exposed to HBV, that person’s immune system may still be primed to mount an immune response sufficient to prevent serious disease. One study of healthcare workers at the University of Wisconsin Hospitals and Clinics found that only 51% of previously vaccinated individuals tested positive for anti-HBs.^8^ However, 89% of anti-HBs negative individuals mounted an anamnestic response when challenged with an HBV booster dose.

This study’s objectives are to present misclassification corrected estimates of HBV immune prevalence among adults in the United States and to compare those estimates to the prevalence of self-reported vaccination and anti-HBs positivity. These objectives are pursued using publicly available, routinely collected national survey data so that our findings and approach may be more easily integrated into existing and future HBV prevention research. Immune prevalence rather than vaccination prevalence was chosen as our primary statistic of interest for simplicity, but has the added advantage of being a more direct measure of population-level susceptibility to HBV infection.

## Methods

### Data

This study used public use data on adults age 18 and older from the 2015-2016 cycle of the National Health and Nutrition Examination Survey (NHANES).^26^ NHANES is a deidentified, cross-sectional population-based survey of the resident noninstitutionalized population of the United States.^27, 28^ NHANES employs a complex probability sampling design that targets several demographic sub-domains deemed of interest by the National Center for Health Statistics.

In the 2015-2016 cycle, a total of 9,971 individuals completed the interview portion of the NHANES survey. 427 of these individuals were not examined at a Mobile Examination Center (where blood is collected for anti-HBs testing) and were excluded. Additionally, 3,942 individuals were under 18 years old, 451 were missing data on anti-HBs status, and 1 was missing data on country of birth. After excluding these individuals, the final sample size was 5,151.

### Naïve outcome measures

#### Prevalence of serological indicators of immunity

Subjects were classified as immune by serology if they tested positive for anti-HBs and classified as not immune if they tested negative.^2, 19^ NHANES uses VITROS immunodiagnostic systems to measure the concentration of serum anti-HBs in each sample using an enhanced chemiluminescence immunoassay (ECi).^29^

#### Self-reported vaccination

Self-reported vaccination status was determined using survey responses. During the interview portion of NHANES, subjects were asked if they had ever received the 3-dose series of the hepatitis B vaccine. Subject responses were classified by NHANES as: at least 3 doses, less than 3 doses, no doses, refused, or don’t know. Anyone who responded that they had received at least 3 doses of vaccine was classified as positive for self-reported vaccination. All others were classified as negative. This classification approach is consistent with how the Centers for Disease Control and Prevention define hepatitis B vaccination among adults when using self-report data.^30^

### Vaccination Timing

Because anti-HBs as an indicator of immunity may be dependent on the age of initial vaccination (see Figure 1), we chose to assign subjects to one of two groups depending on when in their lifetime they were likely to have been vaccinated. The Advisory Committee on Immunization Practices first recommended universal vaccination against hepatitis B for children starting in 1991.^2^ As individuals vaccinated as infants in 1991 would be 24 or 25 years old by the time the 2015-2016 NHANES data were collected, we chose to classify all subjects age 25 and younger as vaccinated in childhood and subjects older than 25 as vaccinated in adolescence or adulthood.

**Figure 1:**
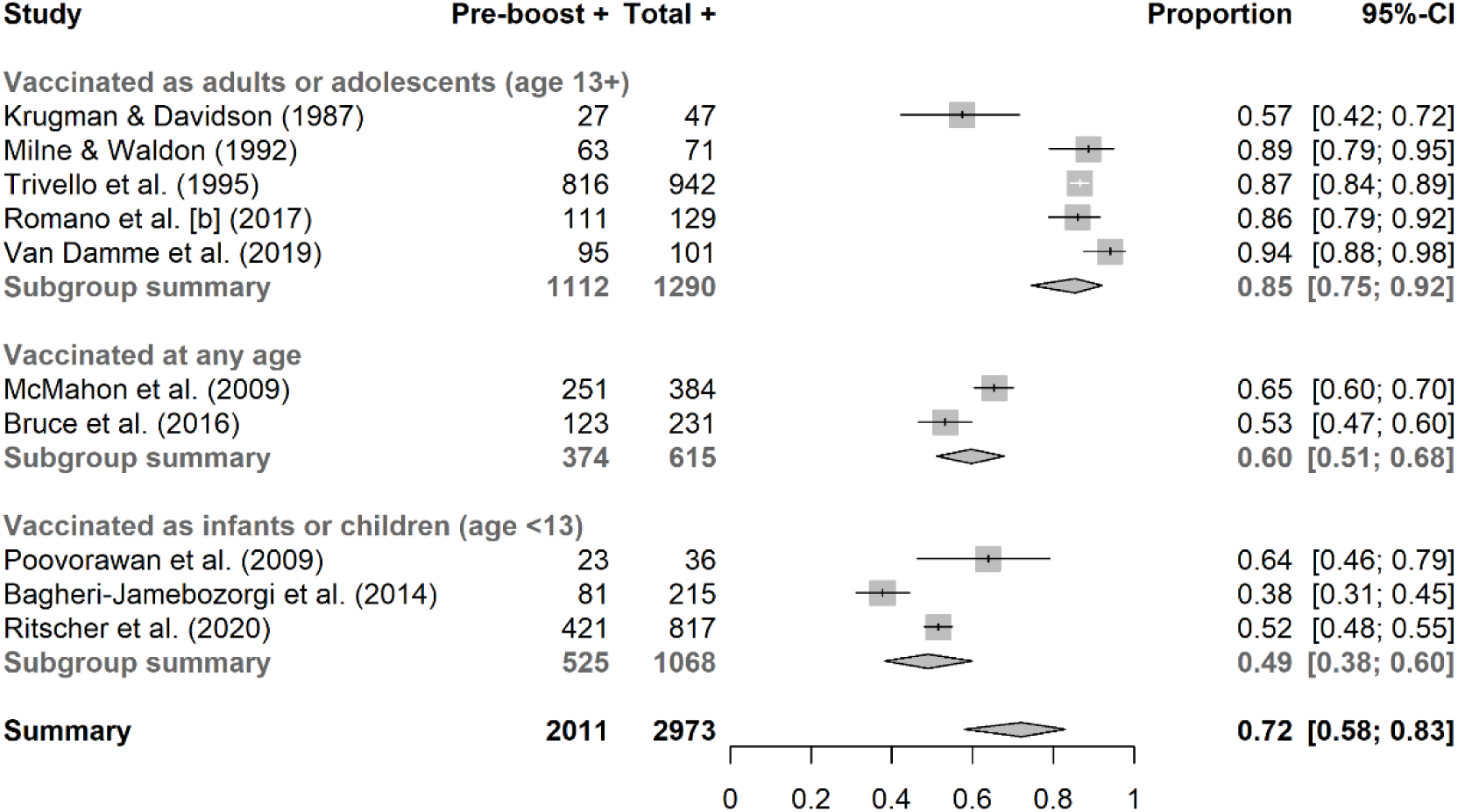
Sensitivity of anti-HBs as a measure of anamnestic response to a booster dose of HBV vaccine among patients who are at least 20 years old. Data from existing literature. Pre-boost+, number of subjects who tested positive prior to receiving the booster shot, adjusting for loss to follow-up; Total +, total number of subjects (pre and post-booster) who tested positive.

### Misclassification correction

We used a Bayesian approach to estimate the prevalence of our three outcome statistics (serological immune prevalence, self-reported vaccination prevalence, and misclassification-corrected HBV immune prevalence), drawing from methods described by Gustafson and implemented by Goldstein et al. and Luta et al. to correct for misclassification.^31–34^ Serological immune status (anti-HBs) was the misclassification-prone measure of immunity that served as the foundation for the misclassification-corrected immune prevalence statistic.

There were three core components to the model we used to calculate misclassification-corrected estimates of immune prevalence, and the first described the relationship between serological immune status and true immune status:

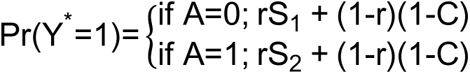

Where Y* is (possibly misclassified) immune status as determined by serological testing, *r* is the probability of truly being immune, *A* indicates the age at primary vaccination, *S_1_* is the sensitivity 5 of the serological test among individuals age 25 and younger, *S_2_* is the sensitivity among individuals age 26 and older, and *C* is the specificity of the serological test. Using two sensitivity values allowed us to differentiate between individuals who were likely vaccinated in childhood and those who were likely vaccinated in adulthood.

The second component of the model described the probability of truly being immune (*r*) given race (Asian American, Black, Hispanic, white, or other), age group (18 to 29, 30 to 49, 50+), birthplace (born in the US, not born in the US), income-poverty ratio (<= 130%, >130%), and self-reported vaccination status. The third model component described the probability of self-reported vaccination using the same covariables.

All coefficients in the second and third model components were given non-informative priors of Normal(0, 0.0001). Choice of appropriate priors for *S* and *C* in the measurement model were a critical component of this analysis, but validation data for anti-HBs as an indicator of HBV immune status was unavailable within NHANES. Therefore, we turned to external sources to inform these prior distributions.

#### Specificity priors

We conceptualized specificity as the ability for the testing instrument to correctly classify subjects who were truly anti-HBs negative as negative (anti-HBs < 10 mIU/mL). For this prior we drew from two studies on the validity of the VITROS anti-HBs diagnostic tool (which was used by NHANES to determine anti-HBs status). One study reported a specificity of 97.9% (46 true negatives and 1 false positive) and the other a specificity of 92.8% (219 true negatives and 17 false positives).^35, 36^ Given a total of 265 true negative and 18 false positive tests across both studies, we set a specificity prior of Beta(265+1, 18+1), which represents the combination of the data from the aforementioned studies and the equivalent of a uniform prior over the range from 0 to 1 – i.e., a Beta(1,1) distribution – and yields a prior centered at 93.6% (95% CI, 90.2% to 95.9%).

#### Sensitivity priors

To inform anti-HBs sensitivity distributions, we drew from literature on long-term immunity among individuals who had previously been vaccinated against HBV. All studies examined individuals who previously completed an HBV primary vaccination series, tested for anti-HBs prior to administering a booster dose of HBV vaccine, and tested for anti-HBs again 2-4 weeks after administering the booster. Post-booster testing was not required for individuals who initially tested positive. Studies were excluded if subjects had received an HBV booster dose prior to the initial anti-HBs assessment.

If there was a shift from anti-HBs negative (< 10 mIU/mL) to anti-HBs positive status (>= 10 mIU/mL) within 2 to 4 weeks of receiving the booster, the initial test was classified as a false negative. Pre-booster positive tests (anti-HBs 10 >= mIU/mL) were considered to be true positives. When there was loss-to-follow-up between the pre- and post-booster test, we multiplied the number of initial positives by the proportion of initial negatives who were not lost-to-follow-up before post-booster test. This adjustment was made to prevent loss to follow-up from artificially inflating sensitivity estimates.

To fully contextualize the sensitivity of anti-HBs as an indicator of immunity, we examined data from any study that met our initial selection criteria and provided relevant data. However, our sensitivity priors were only influenced by studies that examined immunity among previously vaccinated individuals who were at least age 20 or older. In total, we identified 27 studies on hepatitis B immune memory that met our criteria (Table 1). Of these 27 studies, 10 examined immune response among individuals age 20 or older.

**Table 1:**
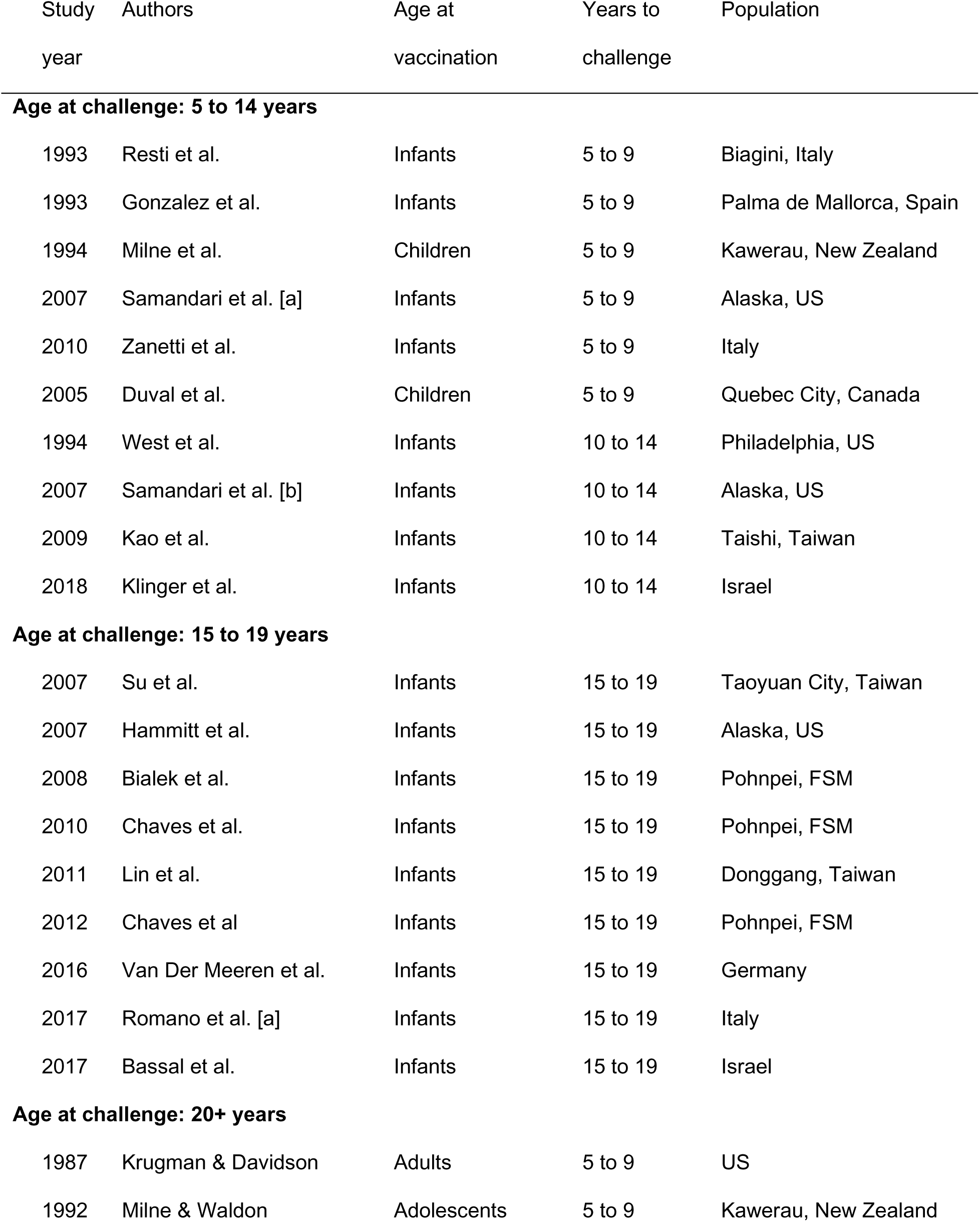
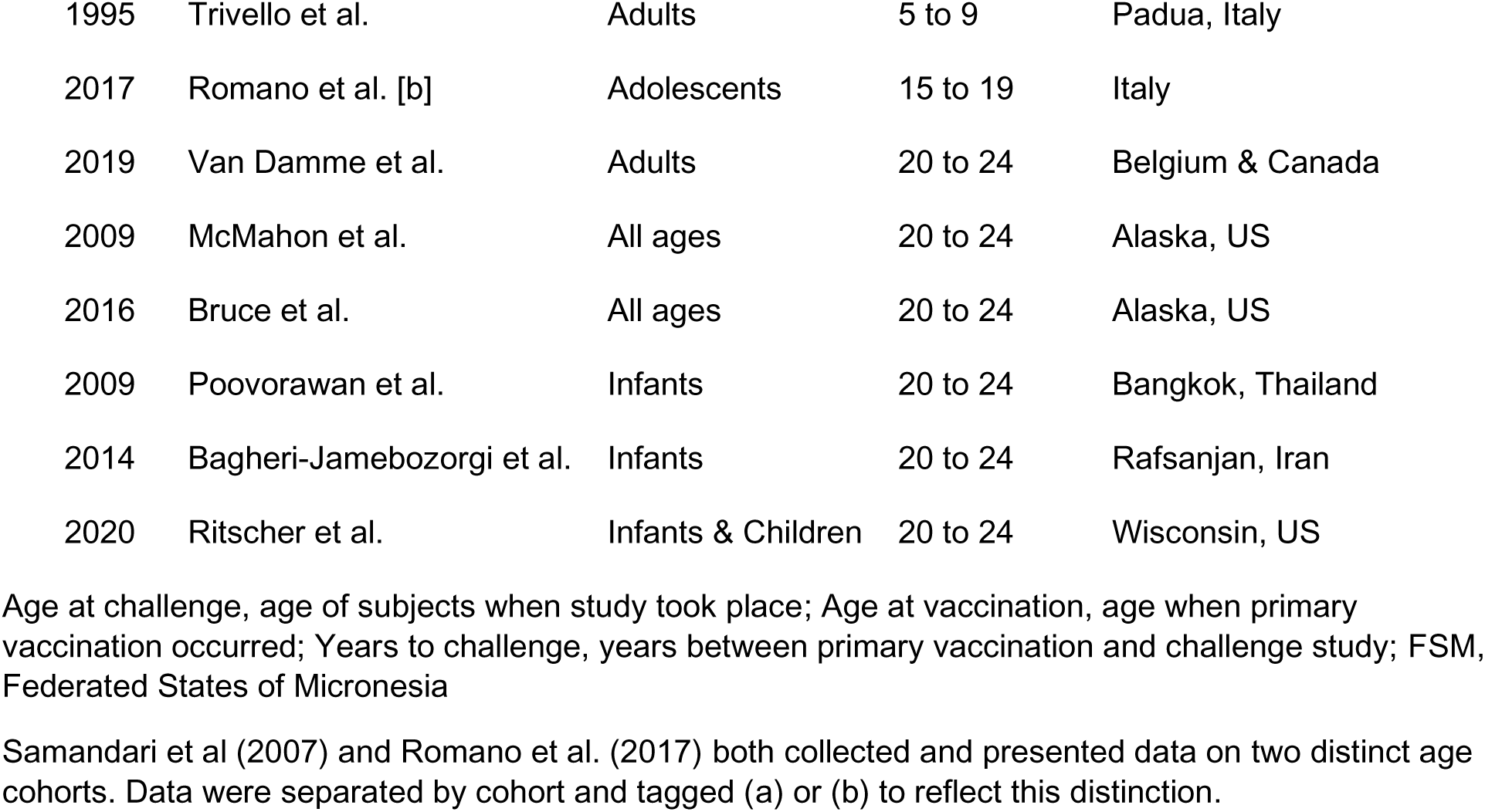
Studies identified to assess sensitivity of anti-HBs as a marker of hepatitis B immunity by number of years post vaccination, age at initial vaccination, and year published.

Figure 1 presents the distribution of pre-booster anti-HBs positivity, post-booster positivity, and estimated sensitivity by study. Five examined post-booster immune response among individuals who received primary vaccination when they were teens (age 13+) or adults, and these had a combined sensitivity of 86.2%. Two studies looked at individuals who received primary vaccination at any age (combined sensitivity of 60.8%). Three studies considered individuals who received primary vaccination in infancy or childhood (age <13), and these studies had a combined sensitivity of 49.2%.

Given the heterogeneity in sensitivity estimates by study, we chose to fit three models with varying sensitivity priors. The specificity prior for each of these models remained the same: 93.6% (95% CI, 90.2% to 95.9%).

In our primary model – Model 1 – the sensitivity prior for subjects age 18 to 25 was set to Beta(106, 110), or 49.1% (42.4% to 55.7%), based on the weighted average sensitivity for individuals who received primary vaccination as infants or children. Sensitivity for subjects age 26 and older was set to Beta(211,35), or 85.8% (81.2% to 89.8%), reflecting the weighted average sensitivity for individuals who received primary vaccination as teens or adults. We chose not to simply use the total number of true positives and false negatives in the beta distributions as we believed those priors would be overly precise.

Model 2 kept the same sensitivity prior for subjects age 26 and older but based the prior for younger subjects on data from Bagheri-Jamebozorgi and colleagues (Beta(82,135)).^37^ At 38.0% (31.5% to 44.3%), this was the lowest study-specific sensitivity for adults who received primary vaccination as infants or children.

Model 3 kept the same sensitivity prior for younger subjects as Model 1, but increased the sensitivity for older subjects to Beta(96,7) based on data from Van Damme et al.^38^. At 93.5% (87.6% – 97.2%) this was the highest sensitivity indicated for individuals who received primary vaccination as adults or adolescents. Sensitivity and specificity priors are summarized by model in eTable 2.

### Population weights

Subdomains of the NHANES sampling strategy included: age (10-year groups starting at 20 years of age), sex, race / ethnicity (Hispanic, non-Hispanic black, non-Hispanic non-black Asian), and income (at or below 130% of the federal poverty level). To take into account this sampling strategy and better describe prevalence at the national level, the conditional posterior probabilities of immunity, anti-HBs positivity, and self-reported vaccination were weighted by age group, sex, race / ethnicity, income group, and foreign birth status using 5-year estimates from the 2018 American Community Survey Public Use Microdata Samples.^39^

The weighting process may be summarized as follows:

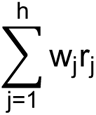

Where w_j_ is the probability of being in demographic stratum *j* in the United States and r_j_ is the posterior probability of immunity, self-reported vaccination, or anti-HBs positivity in stratum *j*. To examine prevalence in demographic subgroups, weights were rescaled by dividing stratum-level population weights by the sum of the stratum-level population weights in the subgroup.

### Software and tools

Bayesian analysis was performed via Markov chain Monte Carlo simulation using a Gibbs sampler. Models were run with 3 chains. Each chain was run for 15,000 iterations, discarding the first 2,000 iterations. All analysis was conducted in R version 4.0.3 (R Foundation for Statistical Computing, Vienna, Austria). The misclassification adjustment model was fit using JAGS version 4.3.0 (http://mcmc-jags.sourceforge.net) and integrated with R using rjags version 4-10. All analytic code may be accessed at https://github.com/daniel-vader/hepb-immune-prev-us/

## Results

### Data

The unweighted distribution of subjects by demographic characteristics, self-reported vaccination status, and anti-HBs status are reported in Table 2. 32.9% of subjects were non-Hispanic white, 31.4% were Hispanic, 20.6% were non-Hispanic Black, 11.4% were non-Hispanic Asian American, and 3.7% did not fall into one of these groups (or belonged to multiple non-Hispanic groups). 48.6% were over 50 years old, 33.1% were 30 to 49 years old, and 18.2% were under 30 years old.

**Table 2:**
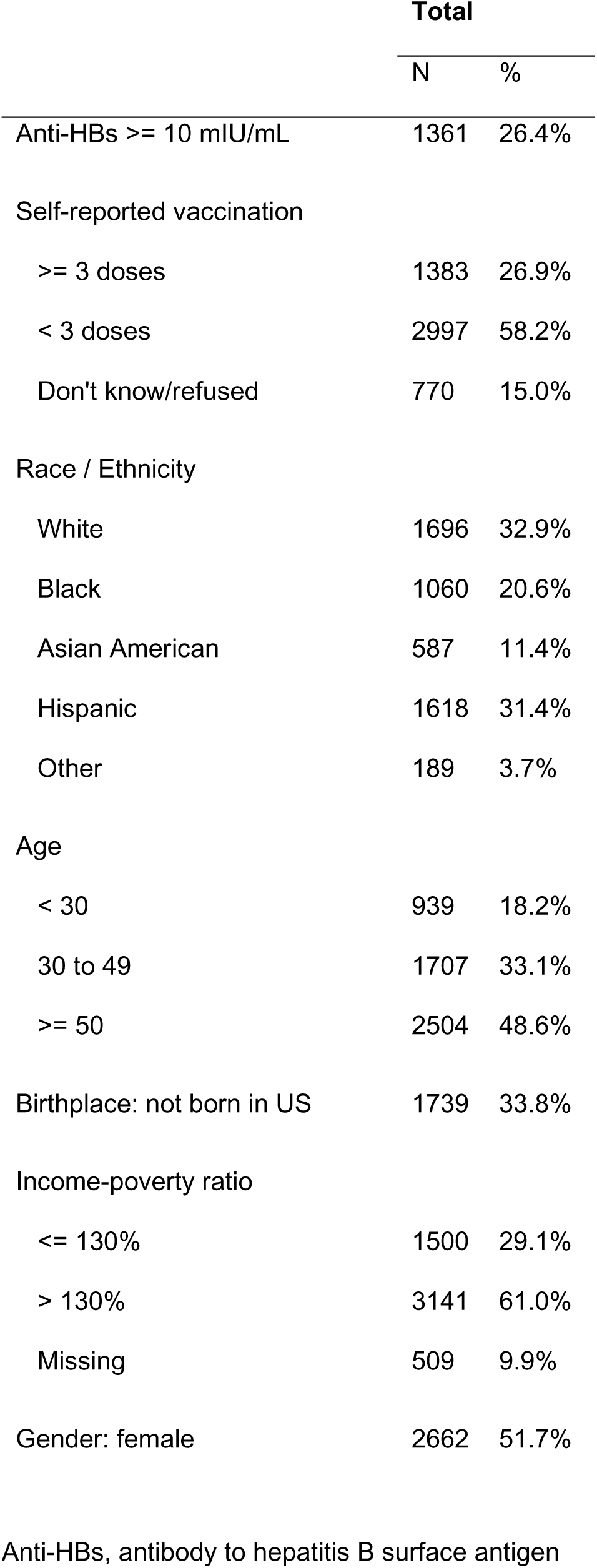
Distribution of sample characteristics. N = 5,151.

|  | Total |  |
| --- | --- | --- |
|  | N | % |
| Anti-HBs ≥ 10 mIU/mL | 1361 | 26.4% |
| Self-reported vaccination |  |  |
| ≥ 3 doses | 1383 | 26.9% |
| < 3 doses | 2997 | 58.2% |
| Don't know/refused | 770 | 15.0% |
| Race / Ethnicity |  |  |
| White | 1696 | 32.9% |
| Black | 1060 | 20.6% |
| Asian American | 587 | 11.4% |
| Hispanic | 1618 | 31.4% |
| Other | 189 | 3.7% |
| Age |  |  |
| < 30 | 939 | 18.2% |
| 30 to 49 | 1707 | 33.1% |
| ≥ 50 | 2504 | 48.6% |
| Birthplace: not born in US | 1739 | 33.8% |
| Income-poverty ratio |  |  |
| ≤ 130% | 1500 | 29.1% |
| > 130% | 3141 | 61.0% |
| Missing | 509 | 9.9% |
| Gender: female | 2662 | 51.7% |

Anti-HBs, antibody to hepatitis B surface antigen

**Table 3:** Estimated prevalence of immunity (bias adjusted), anti-HBs positivity, and self-reported vaccination by demographic group in model 1.

|  | HBV Immunity |  |  | Anti-HBs + |  |  | Self-Reported<br>Vaccination |  |  |
| --- | --- | --- | --- | --- | --- | --- | --- | --- | --- |
|  | Est. | 95% CrI |  | Est. | 95% CrI |  | Est. | 95% CrI |  |
| Marginal | 0.310 | (0.279, | 0.341) | 0.246 | (0.232, | 0.261) | 0.284 | (0.270, | 0.299) |
| Race / Ethnicity |  |  |  |  |  |  |  |  |  |
| White | 0.267 | (0.234, | 0.303) | 0.219 | (0.201, | 0.239) | 0.266 | (0.246, | 0.286) |
| Black | 0.397 | (0.349, | 0.450) | 0.301 | (0.274, | 0.330) | 0.315 | (0.289, | 0.342) |
| Asian | 0.657 | (0.582, | 0.735) | 0.498 | (0.455, | 0.541) | 0.406 | (0.369, | 0.443) |
| Hispanic | 0.273 | (0.231, | 0.315) | 0.213 | (0.192, | 0.235) | 0.281 | (0.258, | 0.305) |
| Other | 0.428 | (0.338, | 0.523) | 0.313 | (0.255, | 0.376) | 0.345 | (0.284, | 0.410) |
| Age |  |  |  |  |  |  |  |  |  |
| 19 to 29 | 0.706 | (0.624, | 0.783) | 0.440 | (0.406, | 0.471) | 0.496 | (0.465, | 0.527) |
| 30 to 49 | 0.247 | (0.208, | 0.288) | 0.227 | (0.204, | 0.252) | 0.322 | (0.299, | 0.347) |
| 50+ | 0.167 | (0.138, | 0.198) | 0.168 | (0.152, | 0.186) | 0.154 | (0.139, | 0.171) |
| Birthplace |  |  |  |  |  |  |  |  |  |
| Born outside US | 0.349 | (0.307, | 0.393) | 0.286 | (0.263, | 0.309) | 0.243 | (0.223, | 0.264) |
| Born within US | 0.302 | (0.271, | 0.334) | 0.239 | (0.223, | 0.255) | 0.292 | (0.276, | 0.309) |

95% CrI, 95% credible interval; HBV Immunity, misclassification-corrected estimate of immunity; anti-HBs +, positive for antibody to hepatitis B surface antigen; self-reported vaccination, reported having received 3 or more doses of hepatitis B vaccine.

### Model fit

Kernel density plots and trace plots for model coefficients and deviance are shown in eFigures 3-8. All models demonstrated reasonable agreement across chains in the density plots and good chain mixing in the trace plots.

### Prevalence estimates

#### Error! Reference source not found

Figure 2 shows the weighted prevalence of immunity, anti-HBs positive serostatus, and self-reported vaccination (at least 3 doses) by demographic group. Table 4 presents numerical prevalence statistics for model 1, while eTable 3 and eTable 4 present statistics for models 2 and 3. In Model 1, an estimated 31.0% (95% credible interval 27.9% to 34.1%) were immune to HBV, 24.6% (23.2% to 26.1%) were anti-HBs positive, and 28.4% (27.0% to 29.9%) reported having received at least 3 doses of HBV vaccine. Variation in the prevalence of HBV immunity was observed by race / ethnicity with the lowest prevalence observed among individuals who were white (26.7%, 23.4% to 30.3%) and highest among individuals who were Asian American (65.7%, 58.2% to 73.5%). Unsurprisingly, estimated prevalence of immunity was highest among 19 to 29-year-olds (70.6%, 62.4% to 78.3%), followed by 30 to 49-year-olds (24.7%, 20.8% to 28.8%), and adults age 50 and over (16.7%, 13.8% to 19.8%). Prevalence of immunity among people born outside of the United States was 34.9% (30.7% to 39.3%) and 30.2% (27.1% to 33.4%) among people born in the United States.

**Figure 2:**
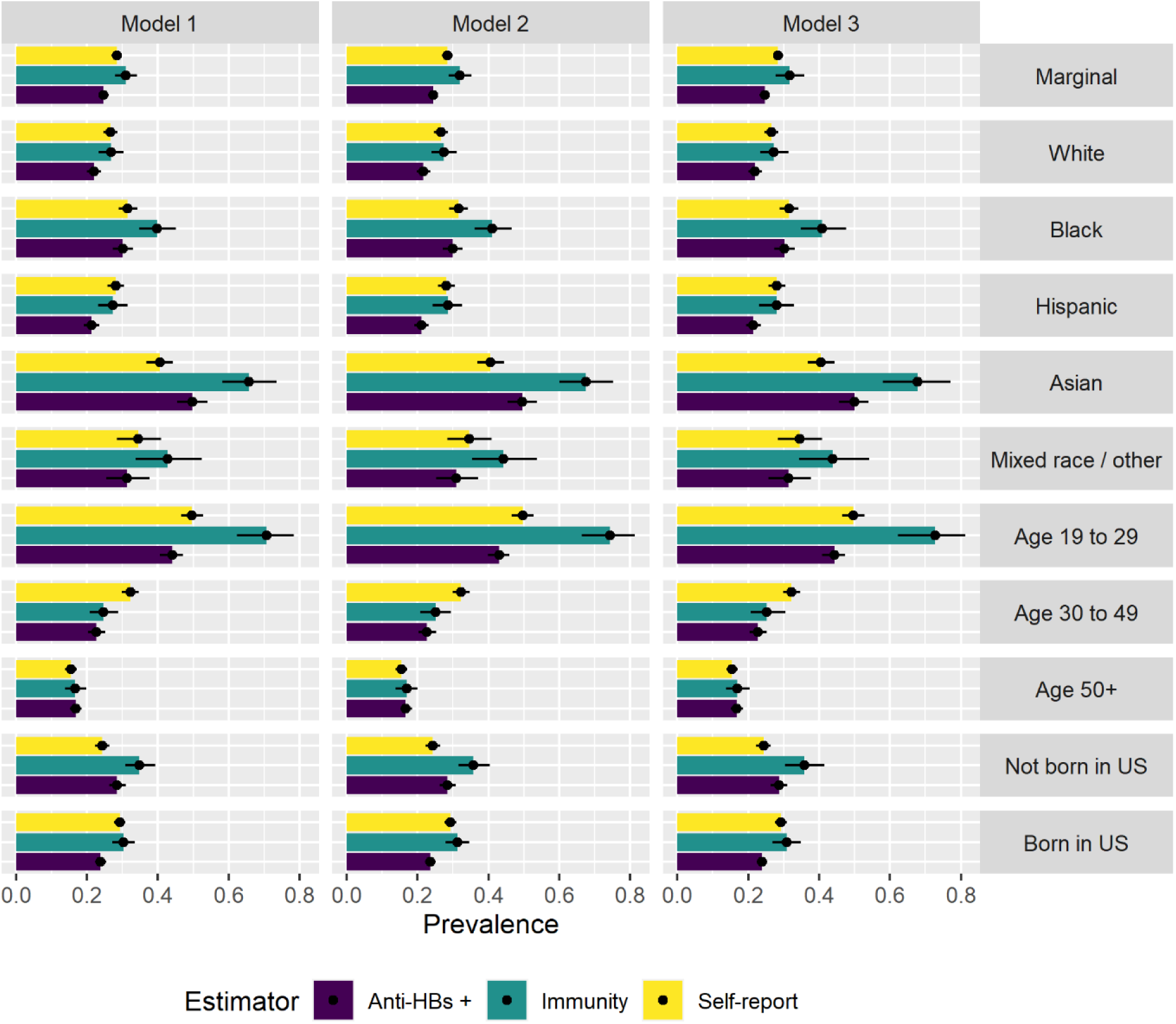
Estimated prevalence of immunity, anti-HBs, and self-reported vaccination by model and demographic group.

**Table 4:** Absolute difference between naïve and bias-corrected estimates of immune prevalence by demographic group in model 1.

|  | Anti-HBs + |  |  | Self-Reported Vaccination |  |  |
| --- | --- | --- | --- | --- | --- | --- |
|  | Est. | 95% CrI |  | Est. | 95% CrI |  |
| Marginal | -0.064 | (-0.088, | -0.039) | -0.026 | (-0.058, | 0.006) |
| Race / Ethnicity |  |  |  |  |  |  |
| White | -0.048 | (-0.071, | -0.025) | -0.002 | (-0.038, | 0.035) |
| Black | -0.096 | (-0.128, | -0.067) | -0.082 | (-0.137, | -0.031) |
| Asian | -0.158 | (-0.209, | -0.115) | -0.251 | (-0.332, | -0.173) |
| Hispanic | -0.060 | (-0.087, | -0.033) | 0.008 | (-0.036, | 0.052) |
| Other | -0.115 | (-0.155, | -0.077) | -0.083 | (-0.183, | 0.015) |
| Age |  |  |  |  |  |  |
| 19 to 29 | -0.265 | (-0.325, | -0.207) | -0.210 | (-0.289, | -0.126) |
| 30 to 49 | -0.020 | (-0.045, | 0.005) | 0.075 | (0.032, | 0.118) |
| 50+ | 0.000 | (-0.020, | 0.023) | -0.013 | (-0.045, | 0.019) |
| Birthplace |  |  |  |  |  |  |
| Born outside US | -0.063 | (-0.093, | -0.036) | -0.106 | (-0.152, | -0.062) |
| Born within US | -0.064 | (-0.088, | -0.040) | -0.010 | (-0.043, | 0.023) |

95% CrI, 95% credible interval; HBV Immunity, misclassification-corrected estimate of immunity; anti-HBs +, positive for antibody to hepatitis B surface antigen; self-reported vaccination, reported having received 3 or more doses of hepatitis B vaccine.

Figure 3 shows the estimated absolute bias that occurred when interpreting serostatus or self-reported vaccination as perfect measures of immunity. In model 1, anti-HBs serostatus underestimated overall prevalence of immunity by 6.4 (3.9 to 8.8) percentage points (Table 4). The degree of bias depended on demographic group, and underestimation was greatest among people who were 19 to 29 years old (26.5%, 20.7% to 32.5%), Asian American (15.8%, 11.5% to 29.9%), or Black (9.6%, 6.7% to 12.8%). Similar patterns were observed in Model 2 (Table S5) and Model 3 (Table S6).

**Figure 3:**
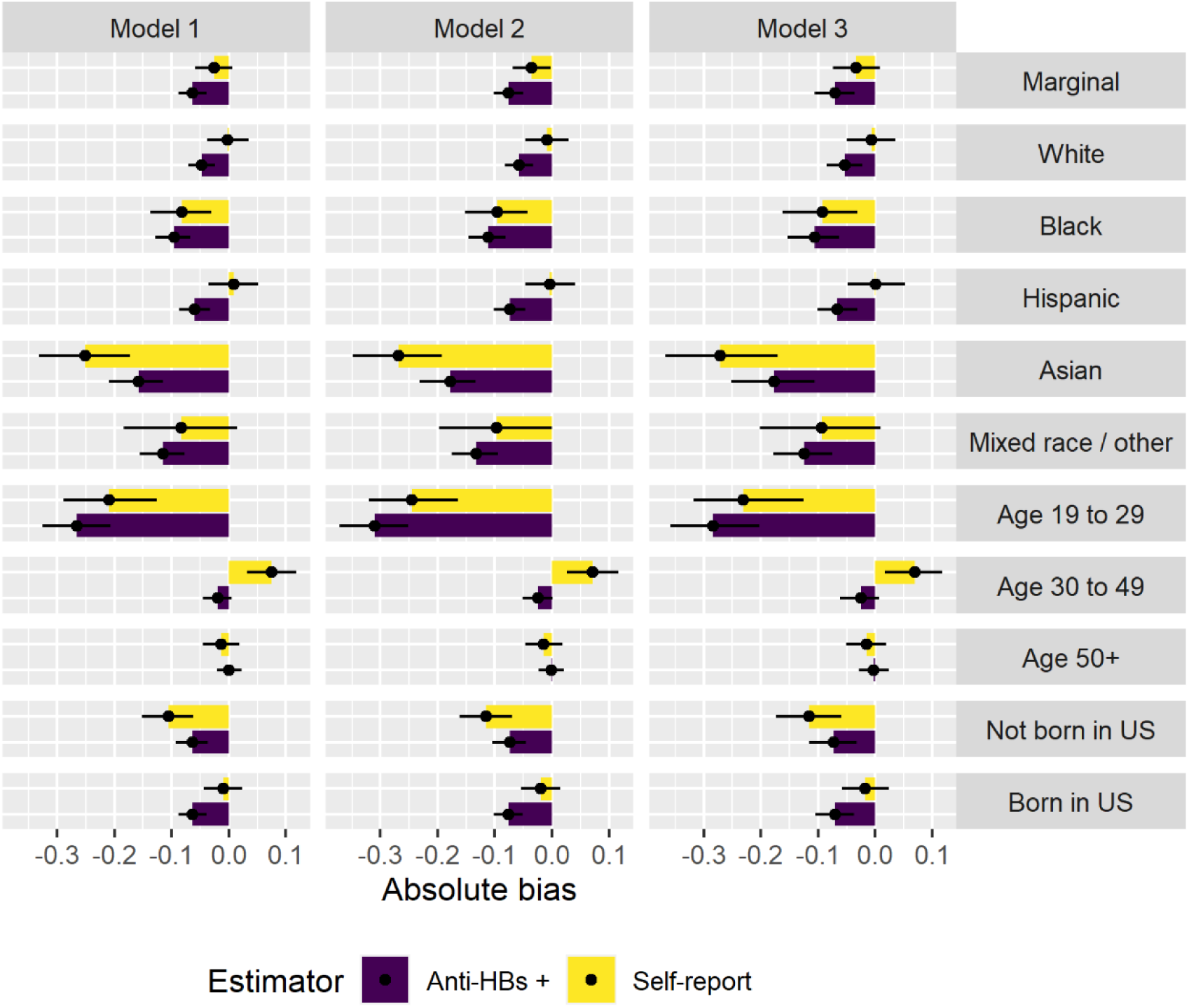
Absolute bias of naïve (anti-HBs or self-report) compared to bias-adjusted estimators of HBV immune prevalence by model and demographic group.

Overall, self-reported vaccination underestimated bias-corrected immunity by 2.6 (−0.6 to 5.8) percentage points. Again, variation was observed by demographic group. The greatest degree of underestimation occurred among people who were Asian American (25.1%, 17.3% to 33.2%), 19 to 29 years old (21.0%, 12.6% to 28.9%), born outside the United States (10.6%, 6.2% to 15.2%), or Black (8.2%, 3.1% to 13.7%). Similar patterns were observed in Model 2 (Table S5) and Model 3 (Table S6). In all models, self-reported vaccination prevalence was higher than immune prevalence in the 30 to 49-year-old age group.

## Discussion

This study estimates that self-reported HBV vaccination prevalence among adults in the US during the 2015-2016 period was 28.4% and that 24.6% of adults were anti-HBs positive. Overall, the estimate for misclassification-corrected immune prevalence (31.0%) was only 2.6 percentage points higher than self-reported vaccination and 6.4 percentage points higher than prevalence of anti-HBs positivity. However, differences were much larger in certain demographic subgroups, particularly people who were 19 to 29 years old, Asian American, or Black.

In 1991, the Advisory Committee on Immunization Practices recommended a strategy for the elimination of HBV transmission in the United States that focused on, among other things, universal childhood vaccination.^2^ In 1995, this recommendation was expanded to recommend vaccination of all unvaccinated 11 and 12-year-olds. In 1999 it was expanded again to include all unvaccinated children under the age of 19.^40^ As these cohorts move farther into adulthood, we expect to see the prevalence of HBV vaccination among young adults to move upward accordingly.

One statistical feature of sensitivity (the probability of testing positive given truly positive) is that its biasing influence on observed probability increases as true prevalence increases. The higher the prevalence of HBV immunity, the greater the influence of sensitivity on estimated immune prevalence. The acute differences observed in this study between bias-corrected immunity, serologically indicated immunity, and self-reported vaccination in the 19 to 25-year-old age group likely reflect the elevated prevalence of vaccination among younger adults compared to older adults. Self-report only performed marginally better than anti-HBs in this age group, which is consistent with the 70.0% sensitivity (and 58.5% specificity) for self-report as an indicator of vaccination estimated by Rolnick and colleagues among 18 to 49-year-olds when using electronic medical records for validation.^41^

NIS Teen estimates of HBV vaccination coverage indicate that about 88% of adolescents between the ages of 13 and 17 were fully vaccinated in 2008.^42^ These 13 to 17-year-olds would be between the ages of 19 and 25 during the 2015-2016 NHANES cycle. Looking at data from 2006, 16 and 17-year-olds (who would be between the ages of 25 and 27 during the 2015-2016 NHANES cycle) had HBV vaccination coverage of 75.6% and 77.3% respectively.^43^ With these statistics in mind, Model 1 estimates of the prevalence of serologically indicated immunity (44.0%) and self-reported vaccination (49.6%) among 19 to 29-year-olds are likely to be underestimations of true HBV immune prevalence. Our approach to misclassification adjustment moved the estimate up to 70.6%, which is closer to what might be expected based on historical data.

When examining literature on anti-HBs as an indicator of HBV immunity, we observed a difference in estimated sensitivity between studies with subjects who were vaccinated as adults or adolescents (where sensitivity was generally higher) and studies with subjects who were vaccinated as infants (where sensitivity was generally lower). This divide may also have consequences for the validity of serologically based estimates of HBV immunity as time since the implementation of universal childhood vaccination in the United States increases.

The sensitivity values chosen for our models were based on studies examining immune memory among previously vaccinated individuals. The second largest study, conducted by Ritscher and colleagues, examined immunity among employees at the University of Wisconsin Hospital who were vaccinated as infants or young children.^8^ It was one of the only studies that examined a non-indigenous North American population. Sensitivity of anti-HBs estimated using data from this study was 52%, which is similar to the sensitivity set in model 1 for individuals likely vaccinated as infants or young children (49.1%).

eFigure 2 presents data on 9 studies on 15 to 19-year-olds. These were not considered when calculating our sensitivity priors but do warrant discussion. Four of these studies indicated unusually low sensitivity values, ranging from 9% to 16%. Of these four, three used the same cohort of subjects from an island in the Federated States of Micronesia.^44–46^ The last study examined immune memory among Alaskan Natives,^47^ and sensitivity estimated from this study’s data was anomalously low compared to three other studies that also looked at immune memory among Alaskan Natives.^19, 25, 48^ These four studies aside, results from Su and colleagues (Taiwan), Lin and colleagues (Taiwan), and Bassal and colleagues (Israel) are consistent with Bagheri-Jamebozorgi et al.’s (Iran) findings as well as findings from other studies on populations from nearby regions.^9, 23, 24, 37, 49^ The sensitivity used for 19 to 29-year-olds in Model 2 (which was based on the Bagheri-Jamebozorgi et al. data) therefore seems reasonable, though perhaps only in groups or regions with greater HBV endemicity.

### Limitations

One important consideration is how time since vaccination affects sensitivity. Though our model takes into account the impact of the timing of initial vaccination on the sensitivity of anti-HBs, it treats sensitivity as constant regardless of the amount of time that has passed since initial vaccination. However, the studies that we do have at hand appear to indicate consistent anti-HBs sensitivity given the amount of time that the HBV vaccine has been in common use. For subjects who were vaccinated as infants or children, the results were relatively consistent if at least 15 years had passed since vaccination. Among individuals vaccinated as adults, data from Van Damme and colleagues suggest high sensitivity up to 20 to 30 years after vaccination.^38^

Another weakness lies in the fact that we cannot perfectly identify who was vaccinated as an infant or child and who was vaccinated later in life. Our approach makes a guess based on the year that someone was born, but there are likely people who were born a year or two prior to 1991 that were still vaccinated when they were very young, and there are likely people born after 1991 who were not vaccinated until they were much older. Failure to incorporate this uncertainty into our model could mean that the estimated credible intervals are too narrow.

## Conclusions

After adjusting for misclassification, we estimate that 31.0% of adults age 19 and older were immune to HBV in the United States in 2015-2016. Using anti-HBs status or self-reported vaccination as surrogates for immunity led to underestimation of immune prevalence, particularly among people who were younger, Asian American, or Black. The consequences of treating these surrogates as unbiased measures of vaccination or immunity may only increase as more vaccinated children age into adulthood.

## Supporting information

eSupplement

## Data Availability

All data are publicly available on the NHANES website.

https://wwwn.cdc.gov/nchs/nhanes/Default.aspx

## Works Cited

1. Centers for Disease Control and Prevention (CDC). Hepatitis B. In: Hamborsky J, Kroger A, Wolfe C, eds. Epidemiology and Prevention of Vaccine-Preventable Diseases. 13th ed. Washington, D.C.: Public Health Foundation; 2015:149–174.

2. Mast EE, Margolis HS, Fiore AE, et al. A Comprehensive Immunization Strategy to Eliminate Transmission of Hepatitis B Virus Infection in the United States Recommendations of the Advisory Committee on Immunization Practices (ACIP) Part 1: Immunization of Infants, Children, and Adolescents. MMWR Recomm Reports. 2005;54(RR-16):1–32. doi:54(RR16)

3. Wasley A, Kruszon-Moran D, Kuhnert W, et al. The Prevalence of Hepatitis B Virus Infection in the United States in the Era of Vaccination. J Infect Dis. 2010;202(2):192–201. doi:10.1086/653622

4. Hepatitis B virus: a comprehensive strategy for eliminating transmission in the United States through universal childhood vaccination. Recommendations of the Immunization Practices Advisory Committee (ACIP). MMWR Recomm reports. 1991;40(RR-13):1–19.

5. Hill HA, Elam-Evans LD, Yankey D, Singleton JA, Kang Y. Vaccination Coverage Among Children Aged 19–35 Months — United States, 2017. MMWR Morb Mortal Wkly Rep. 2018;67:1123–1128. doi:10.15585/mmwr.mm6643a3

6. Centers for Disease Control and Prevention (U.S.). Vaccination Coverage Among Adults in the United States, National Health Interview Survey, 2016. AdultVaxView. https://www.cdc.gov/vaccines/imz-managers/coverage/adultvaxview/pubs-resources/NHIS-2016.html#hepB. Published 2018.

7. Elam-Evans LD, Yankey D, Singleton JA, et al. National, Regional, State, and Selected Local Area Vaccination Coverage Among Adolescents Aged 13–17 Years — United 8. States, 2019. MMWR Morb Mortal Wkly Rep. 2020;69(33):1109–1116. doi:10.15585/mmwr.mm6933a1

8. Ritscher AM, LeClair-Netzel M, Friedlander NJ, et al. Cross-sectional study of hepatitis B antibody status in health care workers immunized as children at an academic medical center in Wisconsin. Vaccine. 2020;38(7):1597–1600. doi:10.1016/j.vaccine.2019.12.036

9. Klinger G, Chodick G, Levy I. Long-term immunity to hepatitis B following vaccination in infancy: Real-world data analysis. Vaccine. 2018;36(17):2288–2292. doi:10.1016/j.vaccine.2018.03.028

10. Krugman S, Davidson M. Hepatitis b vaccine: prospects for duration of immunity. Yale J Biol Med. 1987;60(4):333–338.

11. Trivello R, Chiaramonte M, Ngatchu T, et al. Persistence of anti-HBs antibodies in health care personnel vaccinated with plasma-derived hepatitis B vaccine and response to recombinant DNA HB booster vaccine. Vaccine. 1995;13(2):139–141. doi:10.1016/0264-410X(95)93126-T

12. Milne A, Waldon J. Recombinant DNA Hepatitis B Vaccination in Teenagers: Effect of a Booster at 5½ Years. J Infect Dis. 1992;166(4):942–942. doi:10.1093/infdis/166.4.942

13. Milne A, Hopkirk N, Moyes CD. Hepatitis B vaccination in children: Persistence of immunity at 9 years. J Med Virol. 1994;44(2):113–114. doi:10.1002/jmv.1890440202

14. Bauer T, Jilg W. Hepatitis B surface antigen-specific T and B cell memory in individuals who had lost protective antibodies after hepatitis B vaccination. Vaccine. 2006;24(5):572–577. doi:10.1016/j.vaccine.2005.08.058

15. Resti M, Di Francesco G, Azzari C, Rossi ME, Vierucci A. Anti-HBs and immunological memory to HBV vaccine: implication for booster timing. Vaccine. 1993;11(10):1079. doi:10.1016/0264-410X(93)90145-N

16. Gonzalez ML, Gonzalez JB, Salva F, Lardinois R. A 7-year follow-up of newborns vaccinated against hepatitis B. Vaccine. 1993;11(10):1033–1036. doi:10.1016/0264-410X(93)90129-L

17. Jan CF, Huang KC, Chien YC, et al. Determination of immune memory to hepatitis B vaccination through early booster response in college students. Hepatology. 2010;51(5):1547–1554. doi:10.1002/hep.23543

18. Lu C, Ni Y, Chiang B, et al. Humoral and Cellular Immune Responses to a Hepatitis B Vaccine Booster 15–18 Years after Neonatal Immunization. J Infect Dis. 2008;197(10):1419–1426. doi:10.1086/587695

19. McMahon BJ, Dentinger CM, Bruden D, et al. Antibody Levels and Protection after Hepatitis B Vaccine: Results of a 22-Year Follow-Up Study and Response to a Booster Dose. J Infect Dis. 2009;200(9):1390–1396. doi:10.1086/606119

20. Poovorawan Y, Chongsrisawat V, Theamboonlers A, Bock HL, Leyssen M, Jacquet J-M. Persistence of antibodies and immune memory to hepatitis B vaccine 20 years after infant vaccination in Thailand. Vaccine. 2010;28(3):730–736. doi:10.1016/j.vaccine.2009.10.074

21. Duval B, Gîlca V, Boulianne N, et al. Comparative long term immunogenicity of two recombinant hepatitis B vaccines and the effect of a booster dose given after five years in a low endemicity country. Pediatr Infect Dis J. 2005;24(3):213–218. doi:10.1097/01.inf.0000154329.00361.39

22. Zanetti AR, Romanò L, Giambi C, et al. Hepatitis B immune memory in children primed with hexavalent vaccines and given monovalent booster vaccines: an open-label, randomised, controlled, multicentre study. Lancet Infect Dis. 2010;10(11):755–761. doi:10.1016/S1473-3099(10)70195-X

23. Lin CC, Yang CY, Shih CT, Chen BH, Huang YL. Waning immunity and booster responses in nursing and medical technology students who had received plasma-derived or recombinant hepatitis B vaccine during infancy. Am J Infect Control. 2011;39(5):408–414. doi:10.1016/j.ajic.2010.07.010

24. Kao JT, Wang JH, Hung CH, et al. Long-term efficacy of plasma-derived and recombinant hepatitis B vaccines in a rural township of Central Taiwan. Vaccine. 2009;27(12):1858–1862. doi:10.1016/j.vaccine.2009.01.027

25. Samandari T, Fiore AE, Negus S, et al. Differences in response to a hepatitis B vaccine booster dose among Alaskan children and adolescents vaccinated during infancy. Pediatrics. 2007;120(2). doi:10.1542/peds.2007-0131

26. Centers for Disease Control and Prevention (CDC), National Center for Health Statistics (NCHS). National Health and Nutrition Examination Survey Tutorials. https://wwwn.cdc.gov/nchs/nhanes/tutorials/module3.aspx. Published 2018. Accessed September 10, 2019.

27. Johnson CL, Dohrmann SM, Burt VL, Mohadjer LK. National Health and Nutrition Examination Survey: Sample Design, 2011 – 2014. Vital Heal Stat. 2014;2(162):1–33.

28. Zipf G, Chiappa M, Porter K, Ostchega Y, Lewis B, Dostal J. National Health and Nutrition Examination Survey: Plan and Operations, 1999–2010. Vital Heal Stat. 2013;1(46).

29. Centers for Disease Control and Prevention (CDC), National Center for Health Statistics (NCHS). Natonal Health and Nutrition Examination Survey (NHANES): MEC Laboratory Procedures Manual. Atlanta, GA; 2016.

30. Williams WW, Lu P-J, O’Halloran A, et al. Surveillance of Vaccination Coverage among Adult Populations — United States, 2015. MMWR Surveill Summ. 2017;66(11):1–28. doi:10.15585/mmwr.ss6611a1

31. Gustafson P. Measurement Error and Misclassification in Statistics and Epidemiology: Impacts and Bayesian Adjustments. 1st ed. Boca Raton, FL: Chapman and Hall/CRC; 2004.

32. Goldstein ND, Burstyn I, Welles SL. Bayesian Approaches to Racial Disparities in HIV Risk Estimation among Men Who Have Sex with Men. Epidemiology. 2017;28(2):215–220. doi:10.1097/EDE.0000000000000582

33. Goldstein ND, Welles SL, Burstyn I. To Be or Not to Be: Bayesian Correction for Misclassification of Self-reported Sexual Behaviors among Men Who Have Sex with Men. Epidemiology. 2015;26(5):637–644. doi:10.1097/EDE.0000000000000328

34. Luta G, Ford MB, Bondy M, Shields PG, Stamey JD. Bayesian sensitivity analysis methods to evaluate bias due to misclassification and missing data using informative priors and external validation data. Cancer Epidemiol. 2013;37(2):121–126. doi:10.1016/j.canep.2012.11.006

35. Huzly D, Schenk T, Jilg W, Neumann-Haefelin D. Comparison of nine commercially available assays for quantification of antibody response to hepatitis B virus surface antigen. J Clin Microbiol. 2008;46(4):1298–1306. doi:10.1128/JCM.02430-07

36. Ismail N, Fish GE, Smith MB. Laboratory Evaluation of a Fully Automated Chemiluminescence Immunoassay for Rapid Detection of HBsAg, Antibodies to HBsAg, and Antibodies to Hepatitis C Virus. J Clin Microbiol. 2004;42(2):610–617. doi:10.1128/JCM.42.2.610-617.2004

37. Bagheri-Jamebozorgi M, Keshavarz J, Nemati M, et al. The persistence of anti-HBs antibody and anamnestic response 20 years after primary vaccination with recombinant hepatitis B vaccine at infancy. Hum Vaccines Immunother. 2014;10(12):3731–3736. doi:10.4161/hv.34393

38. Van Damme P, Dionne M, Leroux-Roels G, et al. Persistence of HBsAg-specific antibodies and immune memory two to three decades after hepatitis B vaccination in adults. J Viral Hepat. 2019;26(9):1066–1075. doi:10.1111/jvh.13125

39. U.S. Census Bureau. 2018 American Community Survey 5-year Public Use Microdata Samples. 2020.

40. Centers for Disease Control and Prevention (CDC). Hepatitis B vaccination--United States, 1982-2002. MMWR Morb Mortal Wkly Rep. 2002;51(25):549–552, 563.

41. Rolnick SJ, Parker ED, Nordin JD, et al. Self-report compared to electronic medical record across eight adult vaccines: Do results vary by demographic factors? Vaccine. 2013;31(37):3928–3935. doi:10.1016/j.vaccine.2013.06.041

42. Centers for Disease Control and Prevention (CDC). National, state, and local area vaccination coverage among adolescents aged 13-17 years--United States, 2008. MMWR Morb Mortal Wkly Rep. 2009;58(36):997–1001.

43. Centers for Disease Control and Prevention (CDC). National, state, and local area vaccination coverage among adolescents aged 13-17 years--United States, 2006. MMWR Morb Mortal Wkly Rep. 2007;56(34):885–888.

44. Chaves SS, Groeger J, Helgenberger L, et al. Improved anamnestic response among adolescents boosted with a higher dose of the hepatitis B vaccine. Vaccine. 2010;28(16):2860–2864. doi:10.1016/j.vaccine.2010.01.059

45. Bialek SR, Bower WA, Novak R, et al. Persistence of protection against hepatitis b virus infection among adolescents vaccinated with recombinant hepatitis b vaccine beginning at birth: A 15-year follow-up study. Pediatr Infect Dis J. 2008;27(10):881–885. doi:10.1097/INF.0b013e31817702ba

46. Chaves SS, Fischer G, Groeger J, et al. Persistence of long-term immunity to hepatitis B among adolescents immunized at birth. Vaccine. 2012;30(9):1644–1649. doi:10.1016/j.vaccine.2011.12.106

47. Hammitt LL, Hennessy TW, Fiore AE, et al. Hepatitis B immunity in children vaccinated with recombinant hepatitis B vaccine beginning at birth: A follow-up study at 15 years. Vaccine. 2007;25(39-40):6958–6964. doi:10.1016/j.vaccine.2007.06.059

48. Bruce MG, Bruden D, Hurlburt D, et al. Antibody Levels and Protection after Hepatitis B Vaccine: Results of a 30-Year Follow-up Study and Response to a Booster Dose. J Infect Dis. 2016;214(1):16–22. doi:10.1093/infdis/jiv748

49. Su FH, Cheng SH, Li CY, et al. Hepatitis B seroprevalence and anamnestic response amongst Taiwanese young adults with full vaccination in infancy, 20 years subsequent to national hepatitis B vaccination. Vaccine. 2007;25(47):8085–8090. doi:10.1016/j.vaccine.2007.09.013

