## Supplementary material for "Re-estimating prevalence of hepatitis B virus immunity among adults in the United States: self-reported vaccination, immunologic markers, and bias correction": eSupplement

#### Estimating sensitivity in studies with loss to follow-up

Studies typically only boosted individuals who initially tested negative. In such cases, we assumed that patients who initially tested positive would have also tested positive after boosting. When there was loss-to-follow-up between initial assessment and assessment after boosting, we applied a multiplied the number of initial positives equal to the proportion of initial negatives that were not lost to follow-up.

This scaling factor may be described using the following equation:

$$p(y^* = 1|y = 1) = \frac{B * s}{B * s + A}$$

Where $p(y^* = 1|y = 1)$ is the probability of being anti-HBs positive given true immune status (Sensitivity), B is the number of subjects who tested positive before boosting, and A is the number of subjects who initially tested negative but who switched to positive after boosting. If no loss-to-follow-up occurred, s would equal 1. If 10% of subjects who initially tested negative were lost to follow-up before boosting, s would equal 0.9.

### JAGS model

```
model {
  for(i in 1:n){
    # outcome model, log odds of hep b immunity given predictors
    vac[i] ~ dbern(p_vac[i])
    logit(p_vac[i])<- b0+b[1]*reth_hisp[i]+
      b[2]*reth_black[i]+b[3]*reth_asian[i]+b[4]*reth_other[i]+b[5]*age1[i]+
      b[6]*age2[i]+b[7]*fborn[i]+b[8]*incpov1[i]+b[9]*incpov2[i]+b[10]*srvac1[i]

    # exposure model, log odds of self-reported vaccination (srvac1) given predictors
    srvac1[i] ~ dbern(p_srvac[i])
    logit(p_srvac[i]) <- a0+a[1]*reth_hisp[i]+
      a[2]*reth_black[i]+a[3]*reth_asian[i]+a[4]*reth_other[i]+a[5]*age1[i]+
      a[6]*age2[i]+a[7]*fborn[i]+a[8]*incpov1[i]+a[9]*incpov2[i]

    # measurement model, hepatitis b immunity (vac) is imperfectly measured
      # by anti-HBs (vac_star)
    vac_star[i] ~ dbern(p_vac_star[i])
    p_vac_star[i] <- age25[i]*(sn0*vac[i]+(1-vac[i])*(1-sp0)) +
                     (1-age25[i])*(sn1*vac[i]+(1-vac[i])*(1-sp1))

  }

  # Outcome model priors
  b0 ~ dnorm(0,.0001)
  for(j in 1:10){
    b[j] ~ dnorm(0,.0001)
  }

  # Exposure model priors
  a0 ~ dnorm(0,.0001)
  # Srvac model priors
  for(k in 1:9){
    a[k] ~ dnorm(0,.0001)
  }

  # Measurement model priors (passed in model call)
  sn0~dbeta(sens_a,sens_b)
  sp0~dbeta(spec_a,spec_b)

  sn1~dbeta(sens1_a,sens1_b)
  sp1~dbeta(spec_a,spec_b) # Same prior dist for this spec
}
```

### Tables and figures

**eTable 1** Interpretation of serological tests for hepatitis B. + indicates a positive result, - indicates a negative result, and a blank space indicates that IgM anti-HBc is not required to determine status. Adapted from Mast et al. (2005) and McMahon (2009).[2,26]

| | Serologic Marker | | | |
|---|---|---|---|---|
| | HBsAg | anti-HBc | IgM anti-HBc | anti-HBs |
| Susceptible | - | - | | - |
| Immune due to natural infection | - | + | | + |
| Immune due to hepatitis B vaccination | - | - | | + |
| Acutely infected | + | + | + | - |
| Chronically infected | + | + | - | - |
| Interpretation unclear | - | + | | - |

HBsAg, Hepatitis B surface antigen; anti-HBc, antibody to hepatitis B core antigen; IgM anti-HBc, IgM antibody to hepatitis B core antigen; anti-HBs, antibody to Hepatitis B surface antigen.

**eTable 2:** Sensitivity and specificity model priors

| | Model 1 | Model 2 | Model 3 |
|---|---|---|---|
| Sensitivity (age <= 25) | Beta(106,110) | Beta(82,135) | Beta(106,110) |
| Sensitivity (age > 25) | Beta(211,35) | Beta(211,35) | Beta(96,7) |
| Specificity | Beta(265,19) | Beta(265,19) | Beta(265,19) |

**eTable 3:** Estimated prevalence of immunity (bias adjusted), anti-HBs positivity, and self-reported vaccination by demographic group in model 2.

| | HBV Immunity | | | Anti-HBs + | | | Self-Reported Vaccination | | |
|---|---|---|---|---|---|---|---|---|---|
| | Est. | 95% CrI | | Est. | 95% CrI | | Est. | 95% CrI | |
| Marginal | 0.319 | (0.288, | 0.352) | 0.244 | (0.230, | 0.258) | 0.284 | (0.270, | 0.299) |
| Race / Ethnicity | | | | | | | | | |
| White | 0.274 | (0.240, | 0.310) | 0.217 | (0.198, | 0.236) | 0.266 | (0.246, | 0.286) |
| Black | 0.411 | (0.361, | 0.465) | 0.299 | (0.272, | 0.328) | 0.315 | (0.289, | 0.342) |
| Asian | 0.675 | (0.600, | 0.752) | 0.496 | (0.453, | 0.537) | 0.406 | (0.369, | 0.444) |
| Hispanic | 0.285 | (0.243, | 0.326) | 0.211 | (0.191, | 0.232) | 0.281 | (0.258, | 0.305) |
| Other | 0.442 | (0.354, | 0.537) | 0.309 | (0.253, | 0.371) | 0.346 | (0.285, | 0.408) |
| Age | | | | | | | | | |
| 19 to 29 | 0.742 | (0.663, | 0.813) | 0.431 | (0.400, | 0.459) | 0.497 | (0.466, | 0.527) |
| 30 to 49 | 0.251 | (0.209, | 0.295) | 0.227 | (0.203, | 0.252) | 0.322 | (0.298, | 0.347) |
| 50+ | 0.169 | (0.139, | 0.200) | 0.167 | (0.151, | 0.185) | 0.154 | (0.139, | 0.171) |
| Birthplace | | | | | | | | | |
| Born outside US | 0.358 | (0.316, | 0.404) | 0.284 | (0.262, | 0.307) | 0.243 | (0.223, | 0.264) |
| Born within US | 0.312 | (0.280, | 0.345) | 0.236 | (0.220, | 0.252) | 0.292 | (0.276, | 0.309) |

**eTable 4:** Estimated prevalence of immunity (bias adjusted), anti-HBs positivity, and self-reported vaccination by demographic group in model 3.

| | HBV Immunity | | | Anti-HBs + | | | Self-Reported Vaccination | | |
|---|---|---|---|---|---|---|---|---|---|
| | Est. | 95% CrI | | Est. | 95% CrI | | Est. | 95% CrI | |
| Marginal | 0.318 | (0.277, | 0.358) | 0.247 | (0.233, | 0.261) | 0.284 | (0.270, | 0.299) |
| Race / Ethnicity | | | | | | | | | |
| White | 0.273 | (0.233, | 0.314) | 0.219 | (0.200, | 0.239) | 0.266 | (0.246, | 0.286) |
| Black | 0.408 | (0.348, | 0.476) | 0.303 | (0.274, | 0.332) | 0.315 | (0.289, | 0.342) |
| Asian | 0.677 | (0.579, | 0.770) | 0.499 | (0.456, | 0.540) | 0.406 | (0.369, | 0.443) |
| Hispanic | 0.281 | (0.231, | 0.328) | 0.214 | (0.193, | 0.236) | 0.281 | (0.258, | 0.306) |
| Other | 0.438 | (0.343, | 0.541) | 0.314 | (0.257, | 0.378) | 0.346 | (0.284, | 0.409) |
| Age | | | | | | | | | |
| 19 to 29 | 0.728 | (0.622, | 0.812) | 0.443 | (0.409, | 0.473) | 0.496 | (0.465, | 0.527) |
| 30 to 49 | 0.253 | (0.208, | 0.305) | 0.227 | (0.204, | 0.253) | 0.322 | (0.299, | 0.347) |
| 50+ | 0.170 | (0.137, | 0.204) | 0.167 | (0.151, | 0.185) | 0.154 | (0.139, | 0.171) |
| Birthplace | | | | | | | | | |
| Born outside US | 0.359 | (0.304, | 0.416) | 0.287 | (0.264, | 0.310) | 0.243 | (0.223, | 0.264) |
| Born within US | 0.309 | (0.269, | 0.349) | 0.239 | (0.223, | 0.255) | 0.292 | (0.276, | 0.309) |

**eTable 5:** Absolute difference between naïve and bias-corrected estimates of immune prevalence by demographic group in model 2.

| | Anti-HBs + | | | Self-Reported Vaccination | | |
|---|---|---|---|---|---|---|
| | Est. | 95% CrI | | Est. | 95% CrI | |
| Marginal | -0.076 | (-0.102, | -0.051) | -0.035 | (-0.069, | -0.003) |
| Race / Ethnicity | | | | | | |
| White | -0.058 | (-0.083, | -0.034) | -0.009 | (-0.046, | 0.029) |
| Black | -0.112 | (-0.146, | -0.082) | -0.096 | (-0.152, | -0.043) |
| Asian | -0.178 | (-0.231, | -0.133) | -0.269 | (-0.348, | -0.192) |
| Hispanic | -0.074 | (-0.102, | -0.046) | -0.004 | (-0.047, | 0.040) |
| Other | -0.133 | (-0.175, | -0.094) | -0.097 | (-0.198, | 0.000) |
| Age | | | | | | |
| 19 to 29 | -0.311 | (-0.371, | -0.252) | -0.245 | (-0.320, | -0.164) |
| 30 to 49 | -0.024 | (-0.051, | 0.002) | 0.071 | (0.026, | 0.116) |
| 50+ | -0.002 | (-0.024, | 0.021) | -0.014 | (-0.047, | 0.018) |
| Birthplace | | | | | | |
| Born outside US | -0.074 | (-0.105, | -0.046) | -0.115 | (-0.162, | -0.071) |
| Born within US | -0.076 | (-0.102, | -0.051) | -0.020 | (-0.054, | 0.014) |

**eTable 6:** Absolute difference between naïve and bias-corrected estimates of immune prevalence by demographic group in model 3.

| | Anti-HBs + | | | Self-Reported Vaccination | | |
|---|---|---|---|---|---|---|
| | Est. | 95% CrI | | Est. | 95% CrI | |
| Marginal | -0.071 | (-0.106, | -0.036) | -0.033 | (-0.074, | 0.008) |
| Race / Ethnicity | | | | | | |
| White | -0.054 | (-0.085, | -0.023) | -0.007 | (-0.049, | 0.035) |
| Black | -0.106 | (-0.154, | -0.063) | -0.093 | (-0.162, | -0.031) |
| Asian | -0.177 | (-0.252, | -0.106) | -0.271 | (-0.367, | -0.171) |
| Hispanic | -0.066 | (-0.101, | -0.030) | 0.001 | (-0.048, | 0.052) |
| Other | -0.124 | (-0.178, | -0.075) | -0.093 | (-0.202, | 0.009) |
| Age | | | | | | |
| 19 to 29 | -0.284 | (-0.358, | -0.202) | -0.231 | (-0.319, | -0.125) |
| 30 to 49 | -0.025 | (-0.062, | 0.006) | 0.070 | (0.017, | 0.117) |
| 50+ | -0.003 | (-0.029, | 0.024) | -0.015 | (-0.051, | 0.019) |
| Birthplace | | | | | | |
| Born outside US | -0.072 | (-0.115, | -0.032) | -0.116 | (-0.173, | -0.060) |
| Born within US | -0.070 | (-0.105, | -0.037) | -0.017 | (-0.058, | 0.024) |

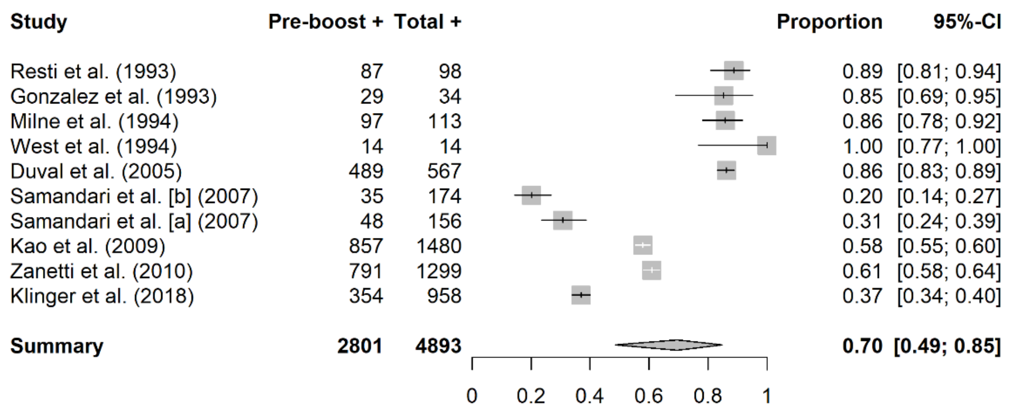


**eFigure 1:** Studies examining anti-HBs positivity pre and post booster 5 to 14 years after completing hepatitis B vaccination during childhood.

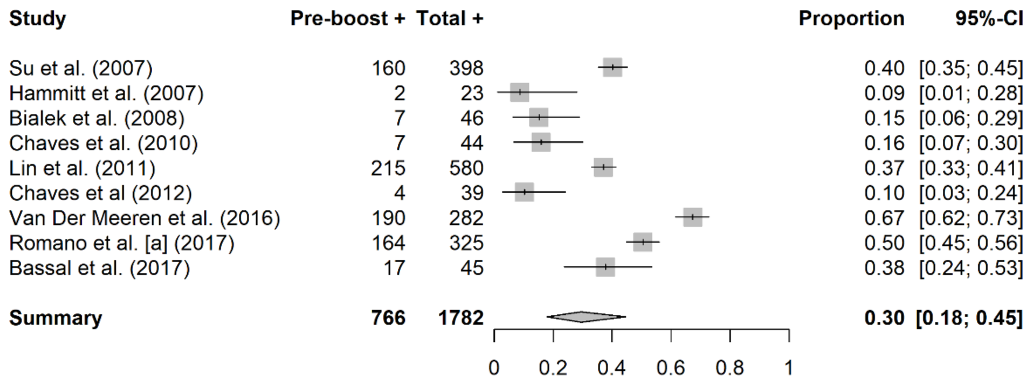


**eFigure 2:** Studies examining anti-HBs positivity pre and post booster 15 to 19 years after completing hepatitis B vaccination during childhood. Note that Bialek et al. (2008), Chaves et al. (2010), and Chaves et al. (2012) examine the same cohort of patients.

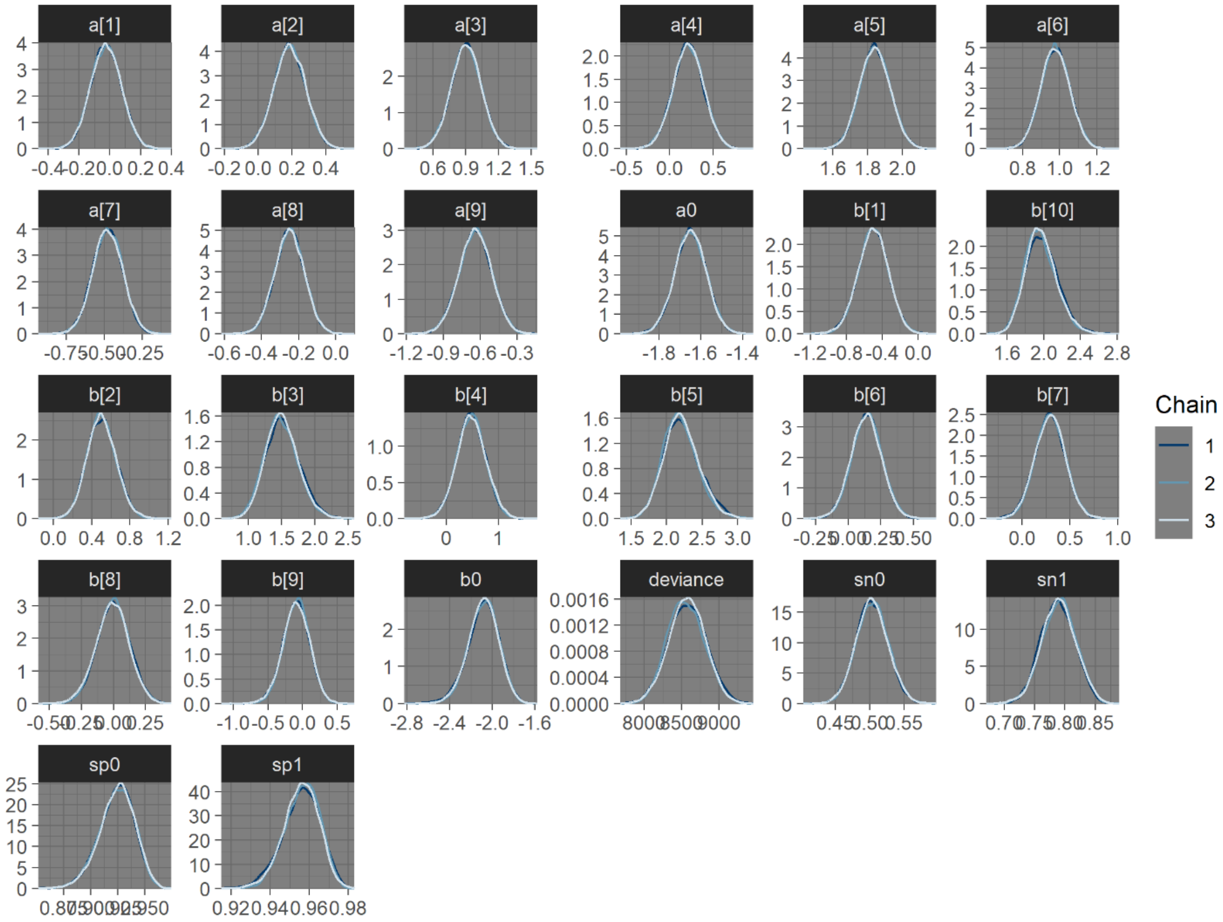


**eFigure 3:** Model 1 kernel density plots for model coefficients and deviance.

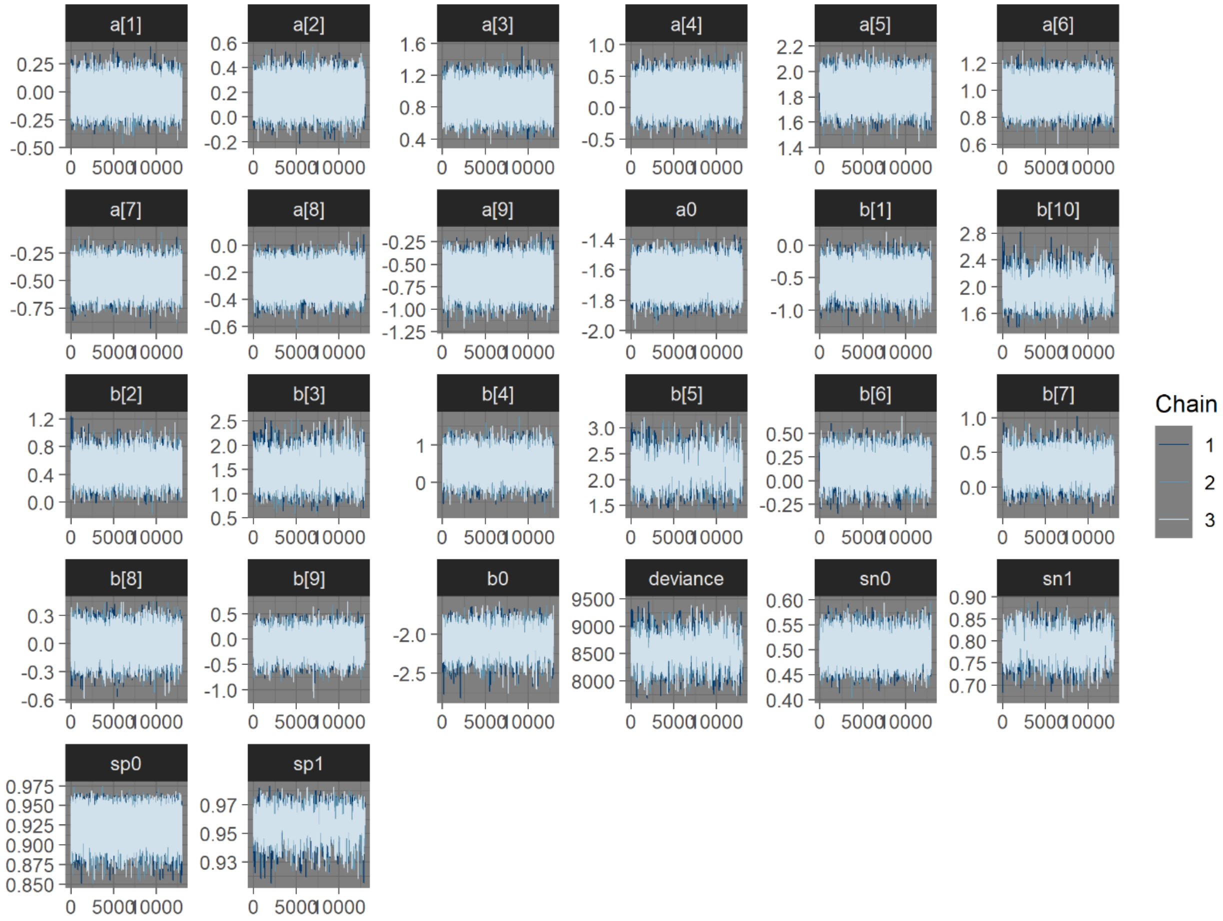


**eFigure 4:** Model 1 trace plots for model coefficients and deviance.

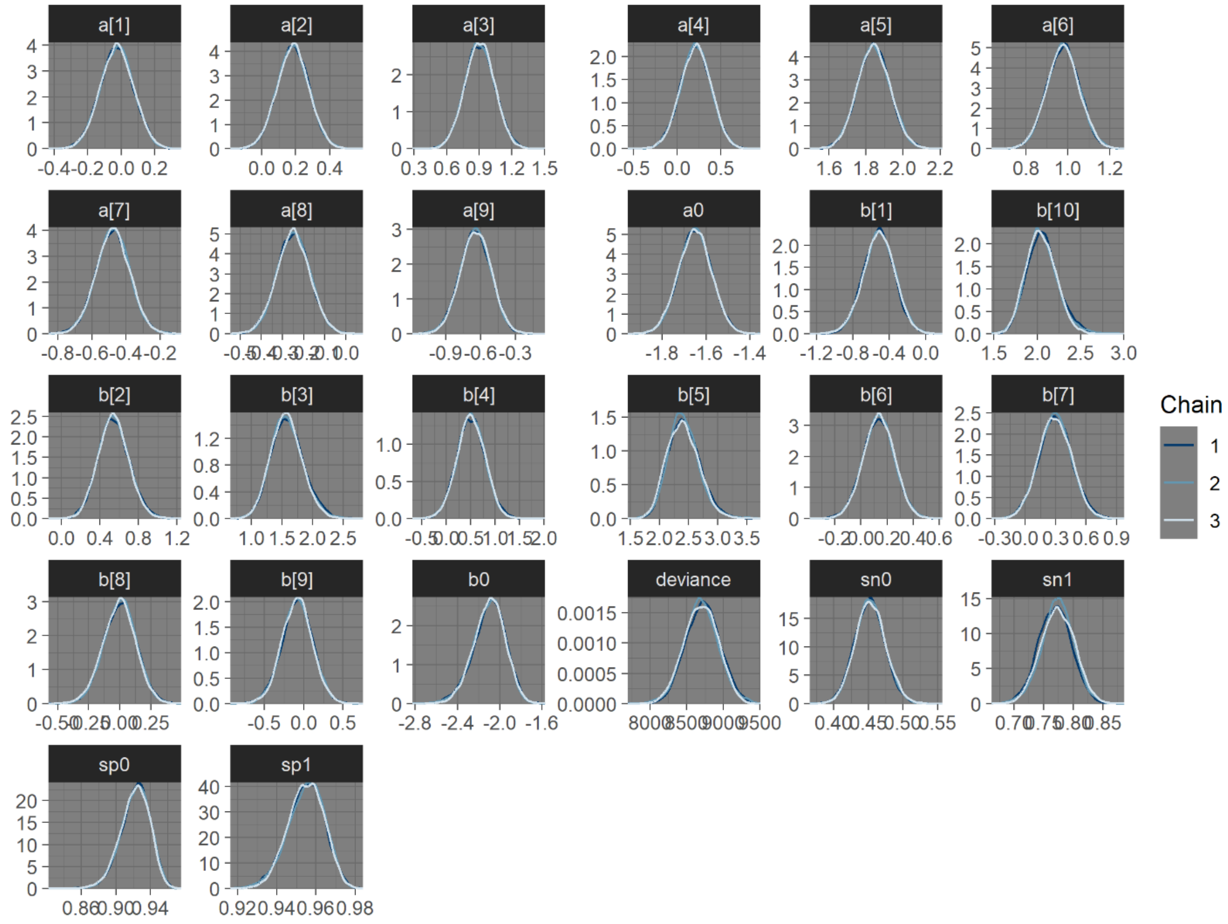


**eFigure 5:** Model 2 kernel density plots for model coefficients and deviance.

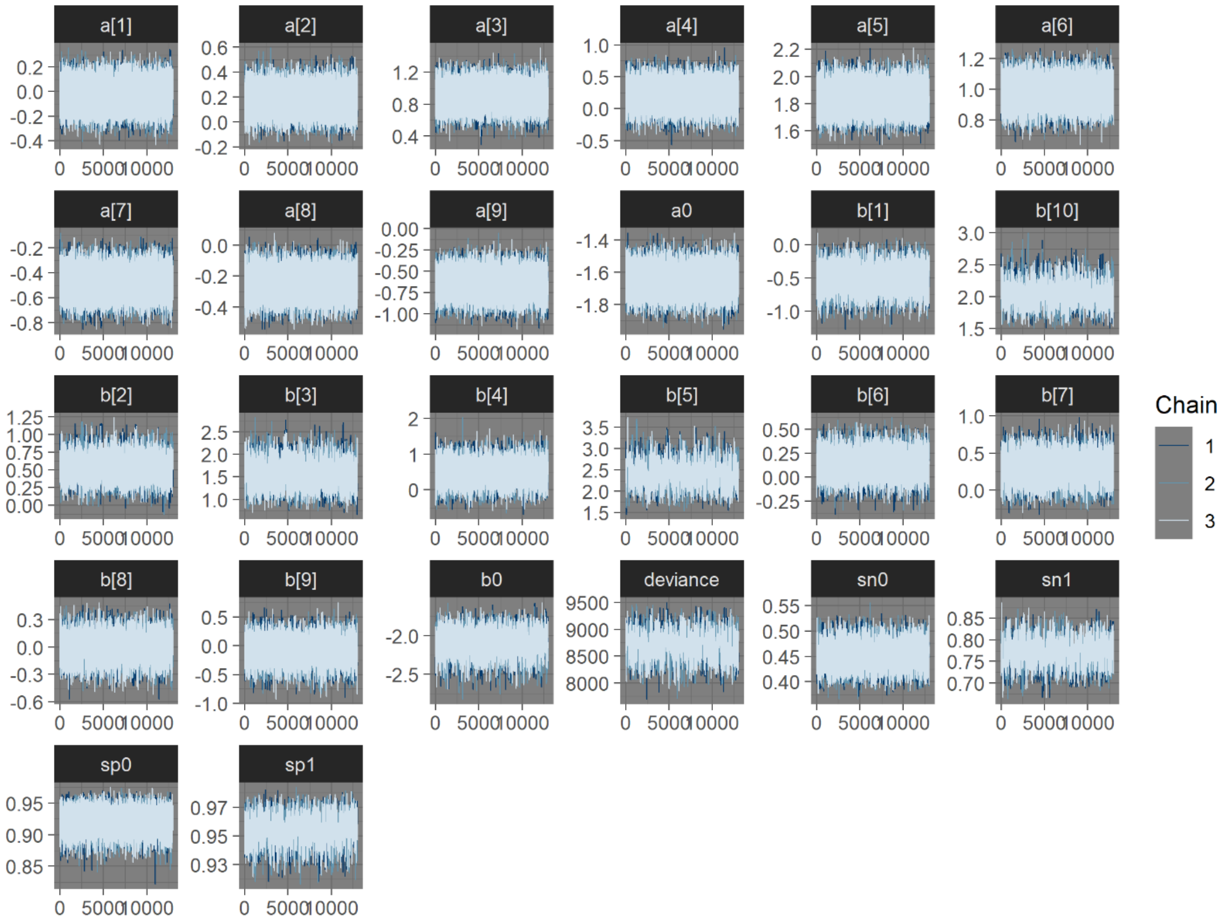


**eFigure 6:** Model 2 trace plots for model coefficients and deviance.

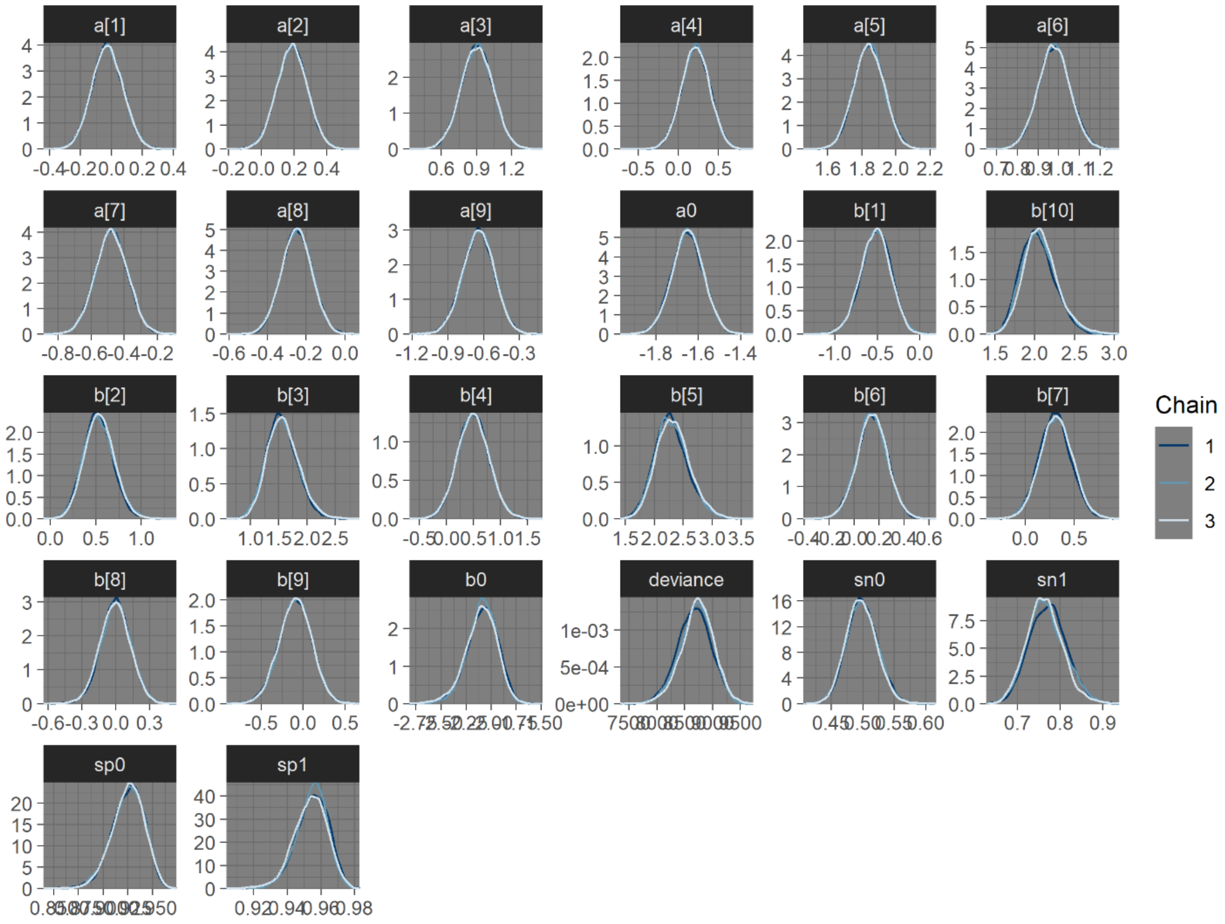


**eFigure 7:** Model 3 kernel density plots for model coefficients and deviance.

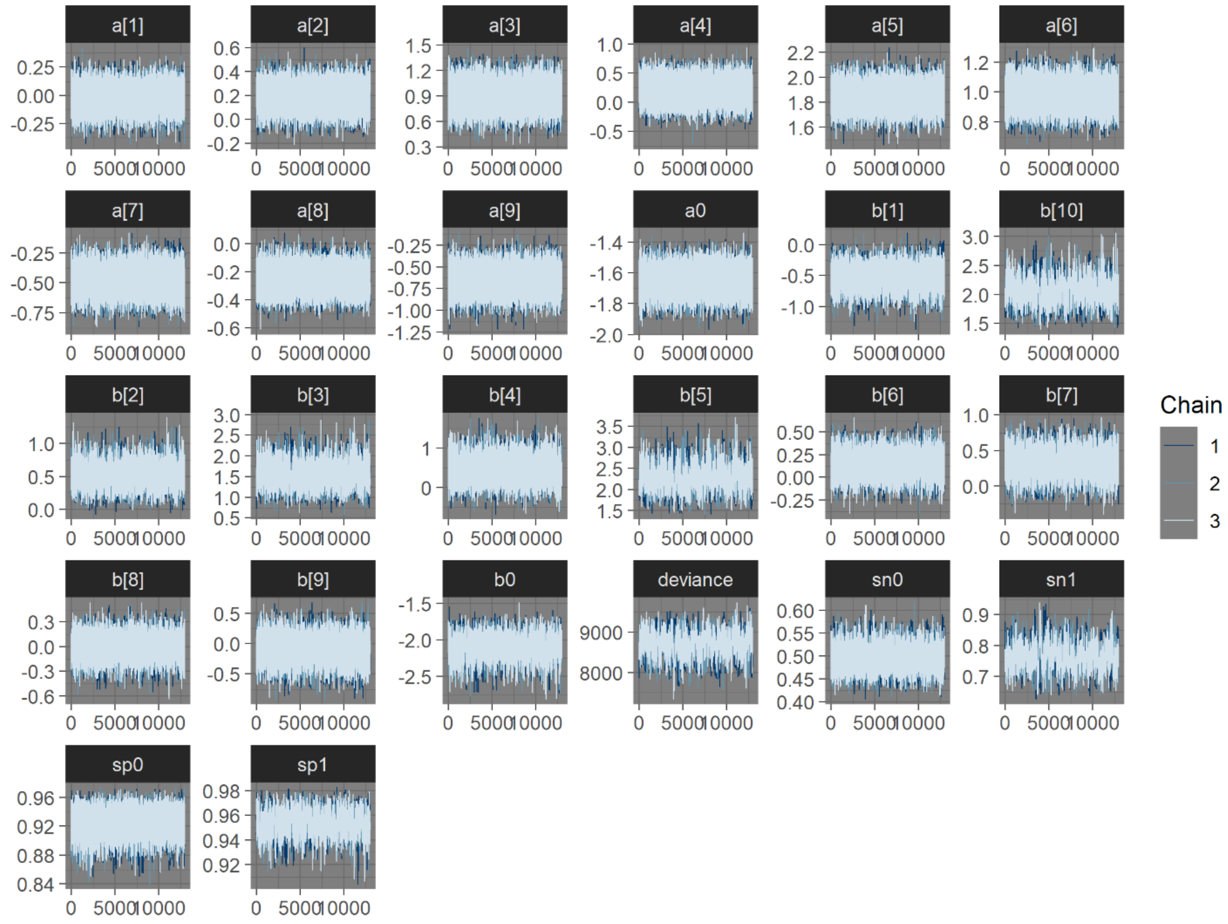


**eFigure 8:** Model 3 trace plots for model coefficients and deviance.
